# Integrating Genome-wide information and Wearable Device Data to Explore the Link of Anxiety and Antidepressants with Heart Rate Variability

**DOI:** 10.1101/2023.08.02.23293170

**Authors:** Eleni Friligkou, Dora Koller, Gita A. Pathak, Edward J. Miller, Rachel Lampert, Murray B. Stein, Renato Polimanti

## Abstract

**Background:** Anxiety disorders are associated with decreased heart rate variability (HRV), but the underlying mechanisms remain elusive.

**Methods:** We selected individuals with whole-genome sequencing, Fitbit, and electronic health record data (N=920; 61,333 data points) from the All of Us Research Program. Anxiety PRS were derived with PRS-CS after meta-analyzing anxiety genome-wide association studies from three major cohorts-UK Biobank, FinnGen, and the Million Veterans Program (N Total =364,550). The standard deviation of average RR intervals (SDANN) was calculated using five-minute average RR intervals over full 24-hour heart rate measurements. Antidepressant exposure was defined as an active antidepressant prescription at the time of the HRV measurement in the EHR. The associations of daily SDANN measurements with the anxiety PRS, antidepressant classes, and antidepressant substances were tested. Participants with lifetime diagnoses of cardiovascular disorders, diabetes mellitus, and major depression were excluded in sensitivity analyses. One-sample Mendelian randomization (MR) was employed to assess potential causal effect of anxiety on SDANN.

**Results:** Anxiety PRS was independently associated with reduced SDANN (beta=-0.08; p=0.003). Of the eight antidepressant medications and four classes tested, venlafaxine (beta=-0.12, p=0.002) and bupropion (beta=-0.071, p=0.01), tricyclic antidepressants (beta=-0.177, p=0.0008), selective serotonin reuptake inhibitors (beta=-0.069; p=0.0008) and serotonin and norepinephrine reuptake inhibitors (beta=-0.16; p=2×10^−6^) were associated with decreased SDANN. One-sample MR indicated an inverse effect of anxiety on SDANN (beta=-2.22, p=0.03).

**Conclusions:** Anxiety and antidepressants are independently associated with decreased HRV, and anxiety appears to exert a causal effect on HRV. Our observational findings provide novel insights into the impact of anxiety on HRV.

## INTRODUCTION

Anxiety disorders are common mental illnesses that affect one out of five patients in a primary care setting(1). In addition to their psychiatric comorbidities, anxiety disorders are associated with multiple somatic diseases(2) consequently leading to increased overall morbidity and mortality(3). Anxiety disorders often co-occur with decreased heart rate variability (HRV)(4), which has been described as an independent predictor of all-cause mortality(5). Decreased HRV can reflect impaired autonomic control(6), a mechanism that has been proposed to contribute to the association of HRV with anxiety(7). Antidepressant medications may also play a role. However, the literature regarding the role of antidepressants in this relationship remains inconclusive, and, in some cases, contradictory(8) The effect of tricyclic antidepressants (TCAs) on HRV(9) has been proposed to account for the decreased HRV in anxiety disorders(10). Conversely, data on the effect of serotonin reuptake inhibitors (SSRIs) are conflicting, with studies showing both increased HRV, in the context of improved depressive symptoms(11), as well as decreased HRV(12). Other factors such as symptom severity and treatment response are likely to contribute to HRV relationship with anxiety and antidepressants.

Because of the multiple potential confounders, it is hard to dissect the underlying mechanisms of the association of anxiety with HRV in traditional observational studies. In recent years, large-scale genome-wide association studies (GWAS) identified a number of variants implicated in the genetic liability to anxiety disorders(13). Since genetic associations are less likely to be affected by reverse causation and other confounders(14), polygenic risk scores (PRS) and genetically informed causal inference analyses can be used to explore complex associations.

To understand the impact of anxiety on autonomic function it is important to measure HRV in real-life settings. Wearable devices are a valuable source of HRV data(15) and full 24-hour HRV metrics can capture circadian variation(16) and response to physical activity(17). In particular, the standard deviation of average five-minute R-R intervals over 24 hours (SDANN) is strongly correlated with other HRV metrics(18) and has been found to be stable over time in the absence of major clinical events, like myocardial infarction(19).

In the present study, we leveraged genome-wide information on anxiety available from UK Biobank (UKB)(20), Million Veteran Program (MVP)(21), and FinnGen(22) and Fitbit data and electronic health records (EHR) from All of Us (AoU) Research Program(23) to investigate the association of anxiety and antidepressants with HRV, and we conducted sensitivity analyses to control for the effect of mental and physical comorbidities.

## METHODS AND MATERIALS

### Study Population

In this retrospective study, we used genetic information, EHRs, physical measurements, and wearable data from participants enrolled in the AoU Research Program(23). When enrolled in the AoU cohort, participants provided informed consent and consent for the release of EHR records separately. Our analyses were conducted from 02/2023 to 04/2023, using the sixth version of the AoU curated data repository (CDR), which was the most recent CDR version as of 02/2023. Age was extracted from AoU self-reported data and baseline body mass index (BMI) from AoU physical measurements. Genomic data were used to derive sex at birth, genetic ancestry, and the first 10 within-ancestry principal components (PCs). Details regarding the quality control of AoU whole-genome sequencing (WGS) data are reported in the **Methods** in **Supplement 1**.

Daily SDANN was calculated for AoU participants that had provided minute-level heart rate Fitbit measurements for at least one full day (1,440 minutes) in a Bring-Your-Own-Device model(24). Fitbit uses photoplethysmography to derive heart rate measurements(25)Data extraction and SDANN calculation were conducted using R (**Methods** in **Supplement 1**). Participants’ health conditions were derived from EHR data that were previously standardized using the Observational Medical Outcomes Partnership (OMOP) vocabulary and SNOMED codes(26). Specifically, we extracted phenotypic information regarding anxiety and conditions that could affect the anxiety-HRV association(27-30). These included myocardial infarction (MI), arrhythmias (Ar), congestive heart failure (CHF), diabetes mellitus (DM) as well as major depressive disorder (MDD). The primary EHR diagnostic subcategories for each phenotype are described in **Table S1**. We also extracted antidepressant prescriptions from EHR data for each date of SDANN measurement (**Table S2**) and assigned them to their respective antidepressant categories (**Table S3**). Our analyses considered antidepressant prescriptions with a start date earlier than the SDANN measurement date and a stop date later than the SDANN measurement date or missing. For our sensitivity analyses, we also accounted for lifetime exposure to beta blockers and calcium channel blockers.

### Polygenic Risk Scoring of Anxiety

To maximize the statistical power of the anxiety PRS analysis, we combined genome-wide association statistics previously generated from three large biobanks: UKB(20), FinnGen(22), and MVP(21). For UKB, we used GWAS data from the Pan-UKB analysis (details at https://pan.ukbb.broadinstitute.org/). Briefly, the genome-wide association analysis was conducted using regression models available in Hail (available at https://github.com/hail-is/hail) and including the top-20 within-ancestry PC, sex, age, age^2^, sex × age, and sex × age^2^ as covariates. For the FinnGen GWAS, we used data from Release 8 (details at https://finngen.gitbook.io/documentation/v/r8/) which were generated with REGENIE(31) including age, sex, top-10 within-ancestry PCs, and genotyping batch as covariates. For both cohorts, we considered definitions of anxiety disorders based on EHR data. Phecode 300 “anxiety disorders” including 10,449 cases and 369,930 controls was used for the UKB GWAS. The FinnGen GWAS was conducted using the endpoint “anxiety disorders” (KRA_PSY_ANXIETY) including 35,385 cases and 254,976 controls. Regarding MVP, we investigated data generated from a previous GWAS of anxiety symptoms assessed via the Generalized Anxiety Disorder (GAD) 2-item scale in 175,163 participants of European descent(32). To meta-analyze GWAS of binary and quantitative phenotypes, we used the sample-weighted approach available in METAL(33), using effective sample size (*N*_*effective*_ = 4/(1/*N*_*cases*_ + 1/*N*_*controls*_)) for the binary phenotypes. The meta-analysis was restricted to GWAS including individuals of European descent because of the limited sample size of other population groups.

Using the anxiety GWAS meta-analysis as input and the 1,000 Genomes Project European populations as linkage disequilibrium (LD) reference, posterior variant-level effect sizes were calculated using PRS-CS(34). Global shrinkage parameter phi was fixed to 1e^-2^. Based on the PRS-CS output, PRS of anxiety in AoU participants was calculated using PLINK 1.9(35).

### Statistical Analysis

All numeric variables investigated in the present study were scaled to have a mean of 0 and a standard deviation of 1.

The association of the anxiety PRS with the daily SDANN measurements was estimated using generalized linear mixed models implemented in the R packages *lme4* and *lmerTest*. Given the multiple observations per person, participant IDs were entered into the model as a random effect grouping factor. The participants’ age in days at the day of the SDANN measurement, BMI at baseline, sex at birth, and the top 10 within-ancestry PCs were included as covariates. To account for the effect of antidepressants on HRV, we include daily antidepressant exposure as an additional covariate in a second model. To verify the robustness of the association, we performed sensitivity analyses excluding individuals with comorbid conditions (i.e., MI, Ar, CHF, DM, and MDD) and medications (beta blockers and calcium channel blockers) that may also affect HRV.

To explore a potential causal effect of anxiety on SDANN, we employed a one-sample Mendelian randomization (MR) approach in AoU(14). The underlying hypothesis of this approach is that if anxiety has a causal effect on SDANN, the anxiety polygenic risk, which is independent of environmental confounders and strongly associated with anxiety, will have a proportional effect on SDANN, provided that there is no other mechanism that links the genetic susceptibility to anxiety and SDANN (horizontal pleiotropy) (**Figure S1**). In that case, we use the anxiety PRS as an instrumental variable to evaluate the relationship of anxiety to SDANN. The analysis was conducted with the R package *ivreg*, using age, sex, BMI, and the top 10 within-ancestry PCs as covariates. The median daily SDANN per person was used as an outcome and lifetime anxiety diagnosis was used as an exposure.

To understand possible differences across antidepressants in their effect on HRV, we used the R packages *lme4* and *lmerTest* to fit generalized linear mixed models of the association of antidepressant use to daily SDANN accounting for age, sex, BMI, anxiety PRS, and the top 10 within-ancestry PCs. The effect of antidepressants on HRV was investigated considering two parallel approaches; testing the effect of specific antidepressant medications (**Table S2**) and the effect of antidepressant categories (**Table S3**). All antidepressant medications were entered simultaneously in one model to estimate their independent effect on HRV. Antidepressant classes were treated accordingly. In the medication-specific analysis, antidepressants prescribed to fewer than ten people were not investigated. We also excluded SDANN measurement dates where the individual had more than four active antidepressant prescriptions, because this may have been due to errors in the EHR registration (e.g., omitted medication end date).

## RESULTS

We analyzed 61,333 data points related to 920 AoU participants of European descent with WGS, Fitbit, and EHR data. Participant baseline characteristics are listed in **Table 1**. The distribution of the daily SDANN values and the number of observations per individual are shown in **Figure 1 and Figure S2** respectively. Due to the diverse number of HRV measurements across AoU participants, we verified the relationship between the number of observations and SDANN estimates (**Methods** in **Supplement 1, Table S4**, and **Figure S2**), observing no statistical association for both daily SDANN (p=0.84) and median SDANN per person (p=0.816).

**Table 1.** Participant baseline characteristics. Demographic characteristics and comorbid conditions of the All of Us Research Program participants

| Participant Characteristic | N (%) / Median (IQR) |
| --- | --- |
| Age at baseline (years) | 51 (24, 79) |
| Sex at birth female | 699 (75) |
| Baseline BMI (kg/m<sup>2</sup>) | 28.9 (8.8) |
| Median SDANN per participant (ms) | 109 (37.5) |
| Anxiety disorder diagnosis | 224 (24) |
| Antidepressant exposed | 219 (24) |
| Major depressive disorder | 208 (23) |
| Congestive heart failure | £ 20 <sup>a</sup> |
| Myocardial infarction | £ 20 <sup>a</sup> |
| Beta blocker or calcium channel blocker | 124 (13) |
| Arrhythmias | 142 (15) |
| Any cardiovascular comorbidity or beta blocker exposure | 213 (23) |
| Diabetes mellitus | 97 (10) |

<sup>a</sup> Participant counts less than 20 not reported due to AoU data protection policy.

**Figure 1.**
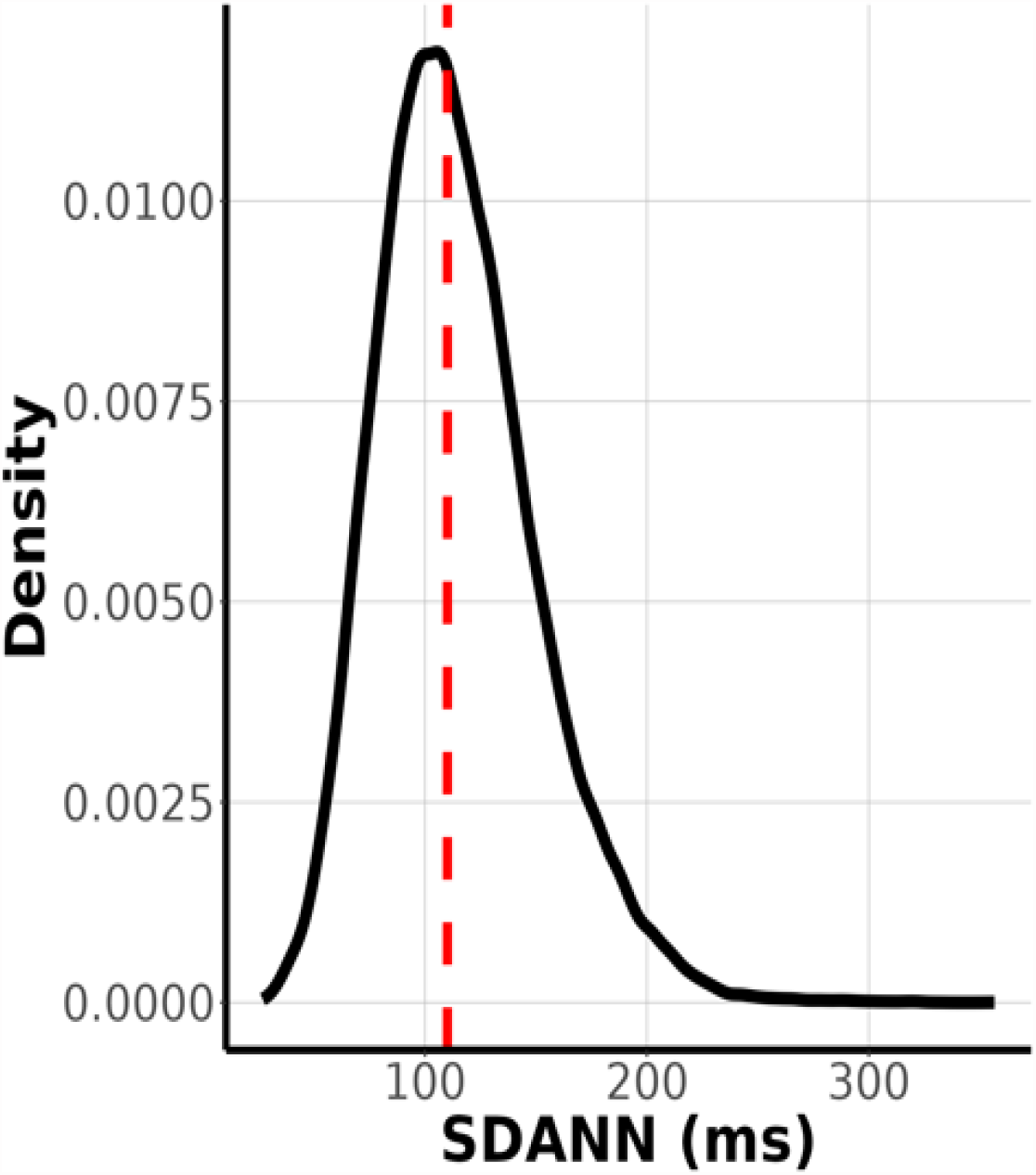
Daily SDANN Values Distribution of Fitbit-derived daily SDANN values for the All of Us Participants

Anxiety PRS was significantly associated with reduced daily SDANN (beta = -0.079, 95%CI= -0.13 – -0.028, p = 0.003) with no changes in the effect after adjusting for antidepressant use in the model [beta = -0.074, 95%CI=-0.13 – -0.027, p = 0.003]. Excluding AoU participants with comorbidities (i.e., MI N £ 20; Ar N= 142; CHF N £ 20; DM N= 97; MDD N= 208) and medications (beta blockers and calcium channel blockers N = 124) that may affect HRV, anxiety PRS remained statistically significantly associated with reduced daily SDANN (**Table S7**). Among the covariates entered in the model, BMI, age, female sex at birth, and antidepressant use were also associated with reduced daily SDANN (**Table S6** and **Figure S3** in Supplement 1).

The one-sample MR analysis demonstrated that the PRS association could indicate a causal effect of anxiety on reduced SDANN (beta=-2.217, p=0.03). The reliability of this estimate is supported by the fact that the anxiety PRS is a robust instrument (P = 0.006). Additionally, SDANN is associated with the variables in our model (Wu-Hausman P = 0.005) and the explanatory variables are significant for the model fit (Wald test P < 0.001) (**Table S5**).

In antidepressant analysis, venlafaxine (beta=-0.12, p=0.002) and bupropion (beta=-0.071, p=0.01) were associated with reduced SDANN. No other antidepressant medications were found to have a significant effect on SDANN (**Figure 2**). When grouping antidepressants by category, TCAs had the strongest effect on SDANN (beta=-0.177; p=8×10^−4^) followed by serotonin and norepinephrine reuptake inhibitors (SNRIs) (beta=-0.157; p=2×10^−6^) and SSRIs (beta=-0.069; p=8×10^−4^). No statistically significant association was observed for serotonin antagonists and reuptake inhibitors (SARIs; beta=0.044, p=0.16; **Figure 3**).

**Figure 2.**
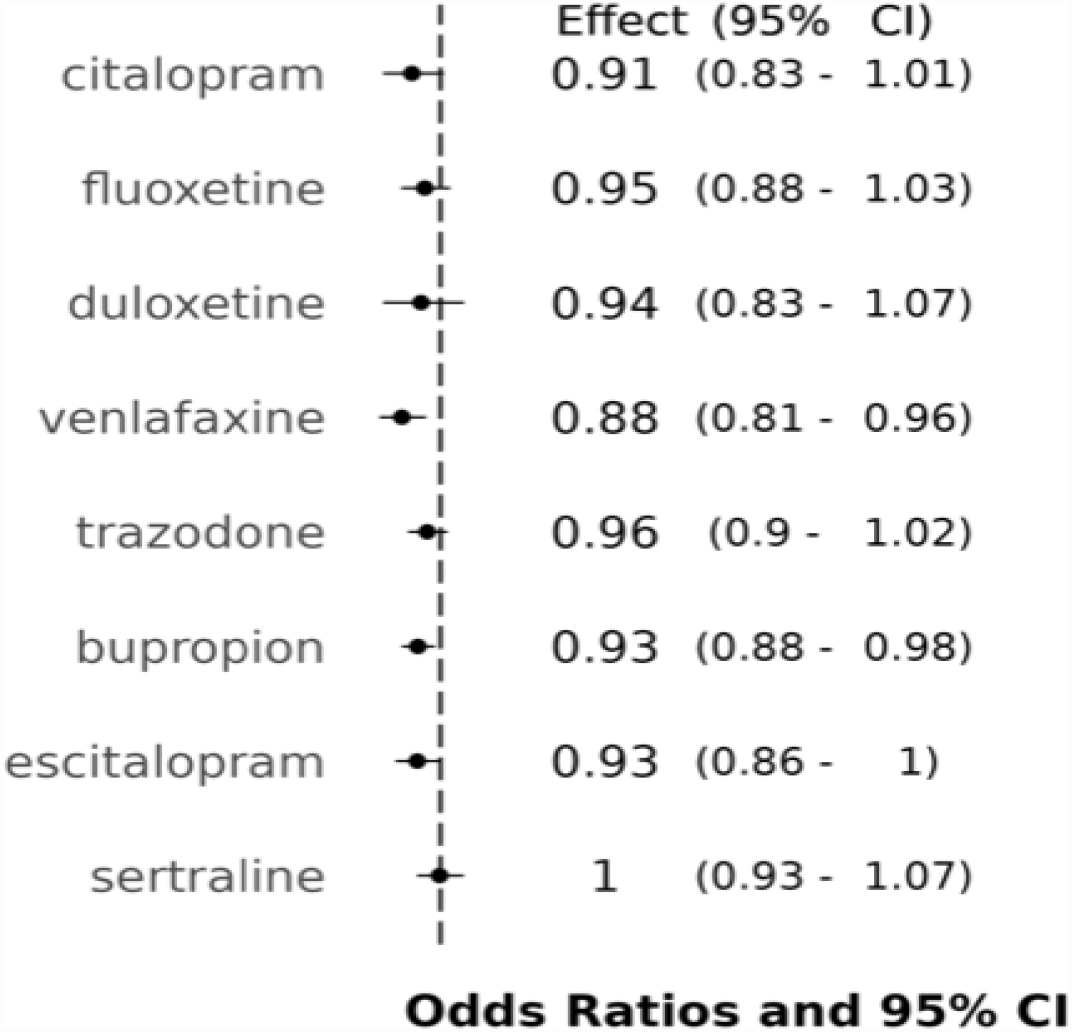
Effect of antidepressant medications on SDANN

**Figure 3.**
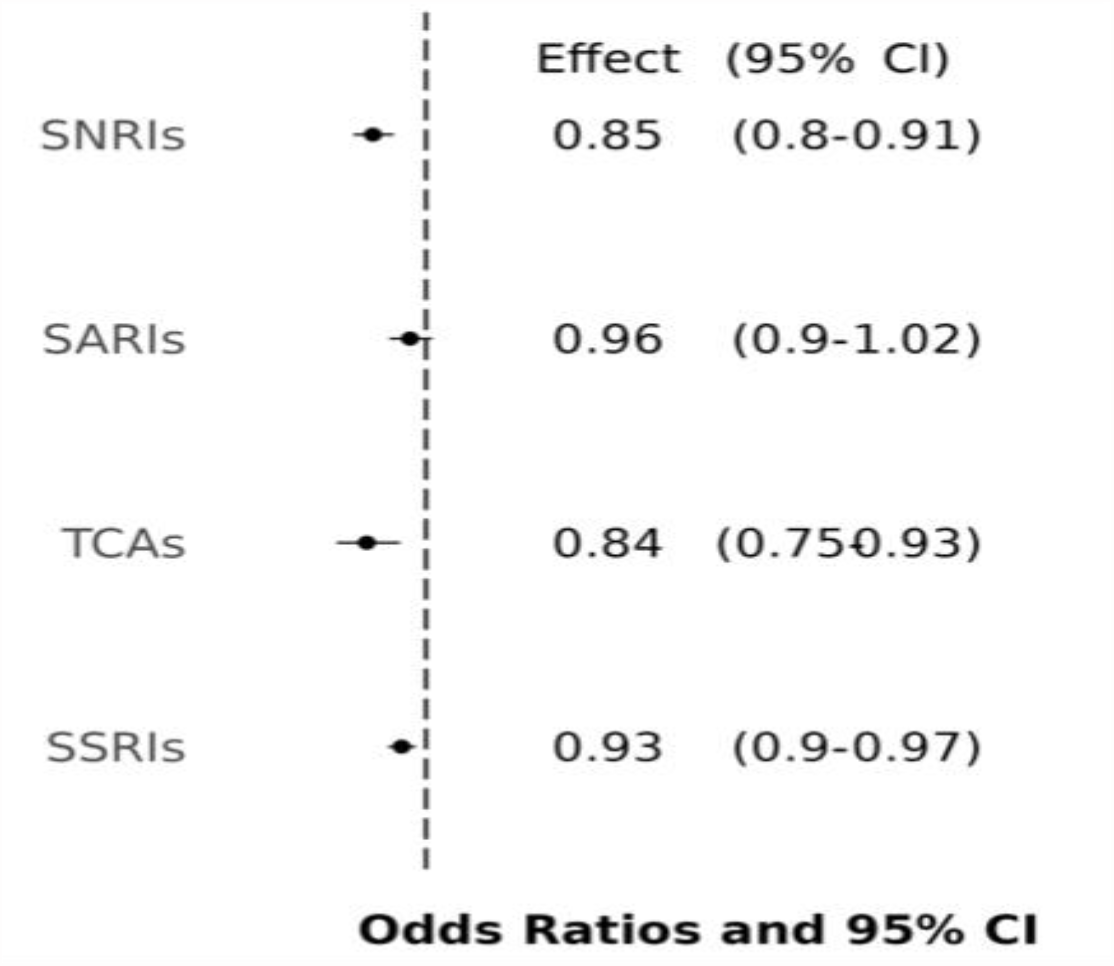
Effect of antidepressant classes on SDANN SSRIs: Selective Serotonin Reuptake Inhibitors; TCAs: Tricyclic Antidepressants; SNRIs: Serotonin and Norepinephrine Reuptake Inhibitors; SARIs: Serotonin Antagonists and Reuptake Inhibitors

## DISCUSSION

Anxiety disorders are a clinically heterogeneous group of mental illnesses with a substantial impact on the global burden of disease that was accentuated following the COVID-19 pandemic(36). Consequently, early identification and intervention are urgently needed. Big data analytics can enrich anxiety diagnostic arsenal and refine existing diagnostic approaches(37). HRV derived by hospital-based measurements has been previously proposed as a predictor of treatment response in individuals with anxious depression(38). To screen for anxiety in the general population, more accessible methods of HRV measurement would be valuable. Digital phenotyping can answer the need for real-life longitudinal HRV measurements prior to the disease onset and identification of a disorder, possibly earning a place in anxiety prediction algorithms(39). Before employing HRV as a predictive tool for anxiety, it is crucial to establish an association independent of comorbid conditions and medications. In this study, we demonstrated that genetic susceptibility to anxiety is associated with decreased SDANN, independently of antidepressant use, cardiovascular comorbidities, DM, and MDD. We additionally provide data supporting a possible causal relationship between anxiety and reduced HRV, supporting the potential use of wearable device-derived SDANN as a predictive tool. To further confirm this, it would be important to test whether the genetic predisposition to reduced HRV may be associated with increased anxiety risk. Unfortunately, at this time it is not possible to perform such analysis; to our knowledge, the only existing HRV GWAS was conducted using HRV measures derived at rest(40). As discussed previously, HRV estimates at rest might reflect distinct aspects of the autonomic function(16, 17).

Beyond dissecting the relationship between HRV and anxiety, we also demonstrated that venlafaxine and bupropion are associated with decreased SDANN independently of anxiety polygenic risk and the use of other antidepressants. These findings support previous evidence; the effect of venlafaxine on HRV was previously reported in a double-blind clinical trial,(41) and bupropion has been associated with decreased HRV in healthy volunteers(42). Given the well-established effect of these drugs on the noradrenergic system(43), our findings are in line with the proposed HRV modulation by brainstem catecholaminergic neurons(43). When we aggregated antidepressants by mechanisms of action, TCAs, SNRIs, and SSRIs were associated with decreased HRV. Both TCAs and SSRIs have been associated with decreased HRV in a recent meta-analysis of randomized placebo-controlled trials (RCTs) and pre-post studies(8). SNRIs have been associated with adverse cardiovascular outcomes, although their effect on HRV as a class has not been investigated to date(44). While our analysis of antidepressant medications expands the knowledge of their effect on HRV, we have to acknowledge the observational nature of our study. Comorbidities and disease severity could simultaneously affect SDANN and drive the clinicians’ choice of antidepressants or the prescribed dosage, potentially confounding the observed effect of the antidepressants on SDANN. Thus, our findings call for further investigation of the role of those medications on HRV using randomized-controlled trials for causal relationships to be established.

Considering the above, the link between HRV, anxiety and antidepressants is complicated by the fact that both disease and treatment affect the outcome in the same direction, making this association challenging to disentangle. The longitudinal 24-hour SDANN values provide a stable metric that supplies the resolution needed in order to investigate variables that change over time, such as antidepressant exposure without requiring hospital-based measurement. Extracting SDANN by minute-level measurements did not allow us to preprocess the inter-beat intervals to remove artifacts or calculate other HRV metrics. Nonetheless, the distribution of SDANN values in our sample is comparable to 24-hour Holter electrocardiogram-derived measures(45-47). Additionally, the anxiety polygenic risk quantifies the genetic predisposition to anxiety in the general population. Of note, our study focused on European individuals, so the generalizability of our findings to other ancestry groups would require further research. As shown in the present study, anxiety polygenic risk is associated with anxiety and antidepressant use, but it is by definition stable over time since birth contrary to the various environmental exposures(48). The significant and robust results of the sensitivity analyses support the association of the anxiety PRS with SDANN, emphasizing the utility and power of genetic instruments in the case of a multifactorial outcome like HRV(49).

In this study, we combined genomic information, wearable device data, and EHRs to investigate the association of HRV with anxiety and antidepressants. Our results provide insights into the impact of anxiety and medications on heart rate variability (HRV), contributing to the study of the comorbidity between mental and physical health. In particular, understanding the mechanisms by which anxiety and antidepressant medications lead to reduced HRV could contribute to a better understanding of the morbidity and mortality observed among individuals affected by mental illnesses.

## Data Availability

Individual patient data are protected by All of Us Research Program Data Protection Policies, and thus cannot be made available.

## ACKNOWLEDGEMENTS

The authors thank the participants and the investigators involved in the UK Biobank, the FinnGen Project, the Million Veteran Program, and the All of Us Research Program. The All of Us Research Program is supported by the National Institutes of Health, Office of the Director: Regional Medical Centers: 1 OT2 OD026549; 1 OT2 OD026554; 1 OT2 OD026557; 1 OT2 OD026556; 1 OT2 OD026550; 1 OT2 OD 026552; 1 OT2 OD026553; 1 OT2 OD026548; 1 OT2 OD026551; 1 OT2 OD026555; IAA #: AOD 16037; Federally Qualified Health Centers: HHSN 263201600085U; Data and Research Center: 5 U2C OD023196; Biobank: 1 U24 OD023121; The Participant Center: U24 OD023176; Participant Technology Systems Center: 1 U24 OD023163; Communications and Engagement: 3 OT2 OD023205; 3 OT2 OD023206; and Community Partners: 1 OT2 OD025277; 3 OT2 OD025315; 1 OT2 OD025337; 1 OT2 OD025276. In addition, the All of Us Research Program would not be possible without the partnership of its participants.

This study was supported by grants from the National Institutes of Health (RF1 MH132337, R33 DA047527, T32 MH014276, and K99 AG078503) and One Mind.

## CONFLICT OF INTEREST STATEMENT

Dr. Polimanti reported receiving personal fees for editorial work on the journal Complex Psychiatry from Karger Publishers and a research grant from Alkermes outside the submitted work. Dr. Stein reported consulting for Acadia Pharmaceuticals Inc, Aptinyx Inc, ATAI Life Sciences, Biogen Inc, Bionomics, BigHealth, BioXcel Therapeutics Inc, Boehringer Ingelheim, Clexio Biosciences Ltd, Eisai Co Ltd, EmpowerPharm, Engrail Therapeutics, Janssen Pharmaceuticals, Jazz Pharmaceuticals, NeuroTrauma Sciences LLC, PureTech Health, Sage Therapeutics, Sumitomo Pharma Co Ltd, and Roche-Genentech; and receiving stock options from Oxeia Biopharmaceuticals Inc and EpiVario outside the submitted work; serving as editor in chief for Depression and Anxiety, deputy editor for Biological Psychiatry, and co–editor in chief for Psychiatry for UpToDate for compensation. Dr. Lampert reported receiving speaker honoraria, advisory board fees, and research funding from Medtronic; speaker honoraria and research funding from Abbott/St Jude; and research funding from Boston Scientific. Dr. Miller reported consulting for Eidos, Pfizer, Siemens, Alnylam, and Roivant and receiving research funding from Eidos, Pfizer, and Argospect. No other disclosures were reported.

## ROLE OF FUNDER/SPONSOR STATEMENT

The funder and sponsors were not involved in design and conduct of the study; collection, management, analysis, and interpretation of the data; preparation, review, or approval of the manuscript; and decision to submit the manuscript for publication.

## Notes

### Author Declarations

All of Us Research Program

## REFERENCES

1. Kroenke K, Spitzer RL, Williams JB, Monahan PO, Löwe B (2007): Anxiety disorders in primary care: prevalence, impairment, comorbidity, and detection. Ann Intern Med. 146:317–325.

2. Härter MC, Conway KP, Merikangas KR (2003): Associations between anxiety disorders and physical illness. Eur Arch Psychiatry Clin Neurosci. 253:313–320.

3. Celano CM, Millstein RA, Bedoya CA, Healy BC, Roest AM, Huffman JC (2015): Association between anxiety and mortality in patients with coronary artery disease: A meta-analysis. Am Heart J. 170:1105–1115.

4. Wang Z, Luo Y, Zhang Y, Chen L, Zou Y, Xiao J, et al. (2023): Heart rate variability in generalized anxiety disorder, major depressive disorder and panic disorder: A network meta-analysis and systematic review. Journal of Affective Disorders. 330:259–266.

5. Filipovic M, Jeger R, Probst C, Girard T, Pfisterer M, Gürke L, et al. (2003): Heart rate variability and cardiac troponin I are incremental and independent predictors of one-year all-cause mortality after major noncardiac surgery in patients at risk of coronary artery disease. Journal of the American College of Cardiology. 42:1767–1776.

6. Bilchick KC, Berger RD (2006): Heart rate variability. Journal of cardiovascular electrophysiology. 17:691–694.

7. Cheng YC, Su MI, Liu CW, Huang YC, Huang WL (2022): Heart rate variability in patients with anxiety disorders: A systematic review and meta-analysis. Psychiatry Clin Neurosci. 76:292–302.

8. Fiani D, Campbell H, Solmi M, Fiedorowicz JG, Calarge CA (2023): Impact of antidepressant use on the autonomic nervous system: A meta-analysis and systematic review. European Neuropsychopharmacology. 71:75–95.

9. Kemp AH, Quintana DS, Gray MA, Felmingham KL, Brown K, Gatt JM (2010): Impact of Depression and Antidepressant Treatment on Heart Rate Variability: A Review and Meta-Analysis. Biological Psychiatry. 67:1067–1074.

10. Hu MX, Milaneschi Y, Lamers F, Nolte IM, Snieder H, Dolan CV, et al. (2019): The association of depression and anxiety with cardiac autonomic activity: The role of confounding effects of antidepressants. Depress Anxiety. 36:1163–1172.

11. Kemp AH, Fráguas R, Brunoni AR, Bittencourt MS, Nunes MA, Dantas EM, et al. (2016): Differential Associations of Specific Selective Serotonin Reuptake Inhibitors With Resting-State Heart Rate and Heart Rate Variability: Implications for Health and Well-Being. Psychosom Med. 78:810–818.

12. Giurgi-Oncu C, Tudoran C, Enatescu VR, Tudoran M, Pop GN, Bredicean C (2020): Evolution of Heart Rate Variability and Heart Rate Turbulence in Patients with Depressive Illness Treated with Selective Serotonin Reuptake Inhibitors. Medicina (Kaunas). 56.

13. Ask H, Cheesman R, Jami ES, Levey DF, Purves KL, Weber H (2021): Genetic contributions to anxiety disorders: where we are and where we are heading. Psychol Med. 51:2231–2246.

14. Davey Smith G, Hemani G (2014): Mendelian randomization: genetic anchors for causal inference in epidemiological studies. Hum Mol Genet. 23:R89–98.

15. Prieto-Avalos G, Cruz-Ramos NA, Alor-Hernández G, Sánchez-Cervantes JL, Rodríguez-Mazahua L, Guarneros-Nolasco LR (2022): Wearable Devices for Physical Monitoring of Heart: A Review. Biosensors (Basel). 12.

16. Frey B, Heinz G, Binder T, Wutte M, Schneider B, Schmidinger H, et al. (1995): Diurnal variation of ventricular response to atrial fibrillation in patients with advanced heart failure. American Heart Journal. 129:58–65.

17. Roach D, Wilson W, Ritchie D, Sheldon R (2004): Dissection of long-range heart rate variability: controlled induction of prognostic measures by activity in the laboratory. J Am Coll Cardiol. 43:2271–2277.

18. Shaffer F, Ginsberg JP (2017): An Overview of Heart Rate Variability Metrics and Norms. Front Public Health. 5:258.

19. Kleiger RE, Bigger JT, Bosner MS, Chung MK, Cook JR, Rolnitzky LM, et al. (1991): Stability over time of variables measuring heart rate variability in normal subjects. Am J Cardiol. 68:626–630.

20. Bycroft C, Freeman C, Petkova D, Band G, Elliott LT, Sharp K, et al. (2018): The UK Biobank resource with deep phenotyping and genomic data. Nature. 562:203–209.

21. Gaziano JM, Concato J, Brophy M, Fiore L, Pyarajan S, Breeling J, et al. (2016): Million Veteran Program: A mega-biobank to study genetic influences on health and disease. J Clin Epidemiol. 70:214–223.

22. Kurki MI, Karjalainen J, Palta P, Sipilä TP, Kristiansson K, Donner KM, et al. (2023): FinnGen provides genetic insights from a well-phenotyped isolated population. Nature. 613:508–518.

23. Denny JC, Rutter JL, Goldstein DB, Philippakis A, Smoller JW, Jenkins G, et al. (2019): The “All of Us” Research Program. N Engl J Med. 381:668–676.

24. Master H, Kouame A, Marginean K, Basford M, Harris P, Holko M (2023): How Fitbit data are being made available to registered researchers in All of Us Research Program. Pac Symp Biocomput. 28:19–30.

25. Natarajan A, Pantelopoulos A, Emir-Farinas H, Natarajan P (2020): Heart rate variability with photoplethysmography in 8 million individuals: a cross-sectional study. Lancet Digit Health. 2:e650–e657.

26. Klann JG, Joss MAH, Embree K, Murphy SN (2019): Data model harmonization for the All Of Us Research Program: Transforming i2b2 data into the OMOP common data model. PLoS One. 14:e0212463.

27. Stein KM, Borer JS, Hochreiter C, Okin PM, Herrold EM, Devereux RB, et al. (1993): Prognostic value and physiological correlates of heart rate variability in chronic severe mitral regurgitation. Circulation. 88:127–135.

28. Huang M, Shah A, Su S, Goldberg J, Lampert RJ, Levantsevych OM, et al. (2018): Association of Depressive Symptoms and Heart Rate Variability in Vietnam War-Era Twins: A Longitudinal Twin Difference Study. JAMA Psychiatry. 75:705–712.

29. Longenecker JC, Zubaid M, Johny KV, Attia AI, Ali J, Rashed W, et al. (2009): Association of low heart rate variability with atherosclerotic cardiovascular disease in hemodialysis patients. Med Princ Pract. 18:85–92.

30. Benichou T, Pereira B, Mermillod M, Tauveron I, Pfabigan D, Maqdasy S, et al. (2018): Heart rate variability in type 2 diabetes mellitus: A systematic review and meta-analysis. PLoS One. 13:e0195166.

31. Mbatchou J, Barnard L, Backman J, Marcketta A, Kosmicki JA, Ziyatdinov A, et al. (2021): Computationally efficient whole-genome regression for quantitative and binary traits. Nat Genet. 53:1097–1103.

32. Levey DF, Gelernter J, Polimanti R, Zhou H, Cheng Z, Aslan M, et al. (2020): Reproducible Genetic Risk Loci for Anxiety: Results From ∼200,000 Participants in the Million Veteran Program. Am J Psychiatry. 177:223–232.

33. Willer CJ, Li Y, Abecasis GR (2010): METAL: fast and efficient meta-analysis of genomewide association scans. Bioinformatics. 26:2190–2191.

34. Ge T, Chen C-Y, Ni Y, Feng Y-CA, Smoller JW (2019): Polygenic prediction via Bayesian regression and continuous shrinkage priors. Nature Communications. 10:1776.

35. Purcell S, Neale B, Todd-Brown K, Thomas L, Ferreira MA, Bender D, et al. (2007): PLINK: a tool set for whole-genome association and population-based linkage analyses. Am J Hum Genet. 81:559–575.

36. Santomauro DF, Mantilla Herrera AM, Shadid J, Zheng P, Ashbaugh C, Pigott DM, et al. (2021): Global prevalence and burden of depressive and anxiety disorders in 204 countries and territories in 2020 due to the COVID-19 pandemic. The Lancet. 398:1700–1712.

37. Onnela J-P, Rauch SL (2016): Harnessing Smartphone-Based Digital Phenotyping to Enhance Behavioral and Mental Health. Neuropsychopharmacology. 41:1691–1696.

38. Kircanski K, Williams LM, Gotlib IH (2019): Heart rate variability as a biomarker of anxious depression response to antidepressant medication. Depress Anxiety. 36:63–71.

39. Jacobson NC, Feng B (2022): Digital phenotyping of generalized anxiety disorder: using artificial intelligence to accurately predict symptom severity using wearable sensors in daily life. Translational Psychiatry. 12:336.

40. Nolte IM, Munoz ML, Tragante V, Amare AT, Jansen R, Vaez A, et al. (2017): Genetic loci associated with heart rate variability and their effects on cardiac disease risk. Nat Commun. 8:15805.

41. Davidson J, Watkins L, Owens M, Krulewicz S, Connor K, Carpenter D, et al. (2005): Effects of paroxetine and venlafaxine XR on heart rate variability in depression. J Clin Psychopharmacol. 25:480–484.

42. Siepmann M, Werner K, Schindler C, Mück-Weymann M, Kirch W (2005): The effects of bupropion on heart rate variability in healthy volunteers. J Clin Psychopharmacol. 25:283–285.

43. Patrone LGA, Capalbo AC, Marques DA, Bícego KC, Gargaglioni LH (2020): An age- and sex-dependent role of catecholaminergic neurons in the control of breathing and hypoxic chemoreflex during postnatal development. Brain Res. 1726:146508.

44. Leong C, Alessi-Severini S, Enns MW, Nie Y, Sareen J, Bolton J, et al. (2017): Cerebrovascular, Cardiovascular, and Mortality Events in New Users of Selective Serotonin Reuptake Inhibitors and Serotonin Norepinephrine Reuptake Inhibitors: A Propensity Score-Matched Population-Based Study. J Clin Psychopharmacol. 37:332–340.

45. Zhao R, Li D, Zuo P, Bai R, Zhou Q, Fan J, et al. (2015): Influences of age, gender, and circadian rhythm on deceleration capacity in subjects without evident heart diseases. Ann Noninvasive Electrocardiol. 20:158–166.

46. Van Hoogenhuyze D, Weinstein N, Martin GJ, Weiss JS, Schaad JW, Sahyouni XN, et al. (1991): Reproducibility and relation to mean heart rate of heart rate variability in normal subjects and in patients with congestive heart failure secondary to coronary artery disease. Am J Cardiol. 68:1668–1676.

47. Geovanini GR, Vasques ER, de Oliveira Alvim R, Mill JG, Andreão RV, Vasques BK, et al. (2020): Age and Sex Differences in Heart Rate Variability and Vagal Specific Patterns – Baependi Heart Study. Global Heart.

48. Smoller JW, Andreassen OA, Edenberg HJ, Faraone SV, Glatt SJ, Kendler KS (2019): Psychiatric genetics and the structure of psychopathology. Mol Psychiatry. 24:409–420.

49. Kalin NH (2022): Polygenic Risk Scores and Genetics in Psychiatry. Am J Psychiatry. 179:781–784.

