## Supplement for "Integrating Genome-wide information and Wearable Device Data to Explore the Link of Anxiety and Antidepressants with Heart Rate Variability"

### Supplementary Online Content

**Methods**
**Table S1.** Subcategories of Participant Diagnoses per Phenotype
**Table S2**. Participant counts per antidepressant drug exposure
**Table S3.** Antidepressant substances included in each antidepressant group
**Table S4.** Effect of number of daily SDANN observations per person on SDANN value
**Table S5.** One-sample Mendelian Randomization diagnostic test values
**Table S6.** Daily SDANN association with anxiety PRS accounting for antidepressant exposure, BMI, age, sex at birth and the first ten principal components
**Table S7.** Anxiety PRS effect on SDANN after excluding people with conditions known to affect heart rate varibility
**Figure S1.** Distribution of number of daily SDANN observations per individual
**Figure S2.** Effect of age, sex, BMI and anxiety PRS accounting for the first 10 principal components

**Methods**

**AOU GENOMIC DATA**

AoU whole genome sequencing (WGS) data is available in vcf files in the AoU research workbench. We first used Hail MatrixTable to keep variants that pass the filters; variants were filtered for low QUAL score (QUALapprox<60 for SNPs) indicating low probability of being a variant, for the absence of high-quality genotypes per site (GQ>=20, DP>=10, and AB>=0.2 for heterozygotes), and for excess heterozygosity (z-score cutoff of -4.5). Multi-allelic variants were split, and only SNPs were extracted. Subsequently, we only kept the loci with minor allele frequency (MAF) higher than 1%. Individuals of European descent were extracted, keeping only the maximal independent unrelated sample, generated by the AoU team using the Hail pc_relate function. Finally, the Hail MatrixTable was converted to bed files.

Information regarding genetically inferred sex at birth and genetic ancestry were previously estimated and was available in the the AoU research workbench. The within-ancestry principal components were calculated from the participants' bfiles using PLINK 1.9. Rsids were assigned to the genomic loci using PLINK 1.9 flag --update-name and the dbSNP hg38 reference panel. The SNPs were pruned with the --indep-pairwise PLINK 1.9 flag at a 50kb window shifted 10 SNPs at each step, with an $r^2$ threshold of 0.1, and minor allele frequency (MAF) greater than 0.05. Using only the pruned loci (--extract <.prune.in file>), we calculated PCs for the individuals of European ancestry using the --pca approx PLINK 2 flag[6].

**SDANN calculation**

Minute heart rate (HR) was extracted from AoU, and five-minute averages were calculated for consecutive five-minute intervals. The average HR was used to calculate the average RR duration of each five-minute interval:

$$Average\ RR = \frac{6000}{Average\ HR}\ ms$$

Subsequently, the standard deviation of all the five-minute RR intervals was calculated, yielding the SDANN value (in ms). One full day started at 00:00 and finished at 23:59. Due to the lack of access to inter-beat intervals (IBIs), we did not evaluate our dataset for non-sinoatrial beats.

### Supplementary References


1. Sudlow C, Gallacher J, Allen N, et al. UK biobank: an open access resource for identifying the causes of a wide range of complex diseases of middle and old age. *PLoS Med*. Mar 2015;12(3):e1001779. doi:10.1371/journal.pmed.1001779
2. Kurki MI, Karjalainen J, Palta P, et al. FinnGen: Unique genetic insights from combining isolated population and national health register data. *medRxiv*. 2022:2022.03.03.22271360. doi:10.1101/2022.03.03.22271360
3. Levey DF, Gelernter J, Polimanti R, et al. Reproducible Genetic Risk Loci for Anxiety: Results From ~200,000 Participants in the Million Veteran Program. *Am J Psychiatry*. Mar 01 2020;177(3):223-232. doi:10.1176/appi.ajp.2019.19030256
4. Willer CJ, Li Y, Abecasis GR. METAL: fast and efficient meta-analysis of genomewide association scans. *Bioinformatics*. Sep 01 2010;26(17):2190-1. doi:10.1093/bioinformatics/btq340
5. Ge T, Chen C-Y, Ni Y, Feng Y-CA, Smoller JW. Polygenic prediction via Bayesian regression and continuous shrinkage priors. *Nature Communications*. 2019/04/16 2019;10(1):1776. doi:10.1038/s41467-019-09718-5
6. Purcell S, Neale B, Todd-Brown K, et al. PLINK: a tool set for whole-genome association and population-based linkage analyses. *Am J Hum Genet*. Sep 2007;81(3):559-75. doi:10.1086/519795

### Table S1. Subcategories of Participant Diagnoses

| Anxiety Disorders[a] |
|---|
| **Standard Concept Name** |
| Acute post-trauma stress state |
| Acute situational disturbance |
| Acute stress disorder |
| Agoraphobia |
| Agoraphobia without history of panic disorder |
| Alcohol-induced anxiety disorder |
| Anxiety disorder |
| Chronic post-traumatic stress disorder |
| Dream anxiety disorder |
| Generalized anxiety disorder |
| Mixed anxiety and depressive disorder |
| Nightmares associated with chronic post-traumatic stress disorder |
| Obsessive-compulsive disorder |
| Organic anxiety disorder |
| Panic disorder |
| Panic disorder with agoraphobia |
| Panic disorder without agoraphobia |
| Phobic disorder |
| Posttraumatic stress disorder |
| Psychoactive substance-induced organic anxiety disorder |
| Separation anxiety disorder of childhood |
| Social phobia |
| **Major Depressive Disorder** |
| **Standard Concept Name** |
| Severe major depression, single episode, without psychotic features |
| Major depression, single episode |
| Depressive disorder |
| Premenstrual dysphoric disorder |
| Mild recurrent major depression |
| Moderate major depression, single episode |
| Single episode of major depression in full remission |
| Severe recurrent major depression without psychotic features |
| Recurrent major depression in partial remission |
| Dysthymia |
| Recurrent major depression in full remission |
| Severe major depression, single episode, with psychotic features |
| Recurrent major depressive episodes |
| Recurrent depression |
| Recurrent major depression in remission |
| Recurrent major depressive episodes, mild |

| |
|---|
| Mild major depression, single episode |
| Moderate recurrent major depression |
| Seasonal affective disorder |
| Recurrent major depression |
| Recurrent major depressive episodes, moderate |
| Postpartum depression |
| Recurrent major depressive episodes, severe, with psychosis |
| Chronic depressive personality disorder |
| Major depression single episode, in partial remission |
| Mixed anxiety and depressive disorder |
| **Myocardial Infarction** |
| **Standard Concept Name** |
| Acute subendocardial infarction |
| Subsequent non-ST segment elevation myocardial infarction |
| Old myocardial infarction |
| Acute non-ST segment elevation myocardial infarction |
| Acute myocardial infarction of inferior wall |
| Acute ST segment elevation myocardial infarction |
| Acute ST segment elevation myocardial infarction due to right coronary artery occlusion |
| Acute myocardial infarction of inferolateral wall |
| Acute myocardial infarction of inferoposterior wall |
| Acute myocardial infarction |
| Acute myocardial infarction of anterolateral wall |
| **Congestive Heart Failure** |
| **Standard Concept Name** |
| Congestive heart failure |
| Hypertensive heart disease with congestive heart failure |
| **Congestive Heart Failure** |
| **Standard Concept Name** |
| Chronic congestive heart failure |
| Hypertensive heart and renal disease with (congestive) heart failure |
| **Diabetes Mellitus** |
| **Standard Concept Name** |
| Type 2 diabetes mellitus without complication |
| Type 1 diabetes mellitus without complication |
| Diabetes mellitus without complication |
| Diabetes mellitus |
| Type 2 diabetes mellitus |
| Peripheral circulatory disorder due to type 2 diabetes mellitus |
| Pre-existing diabetes mellitus in mother complicating childbirth |
| Drug-induced diabetes mellitus |
| Gestational diabetes mellitus |
| Type 1 diabetes mellitus |

| |
|---|
| Gestational diabetes mellitus in childbirth |
| Secondary diabetes mellitus |
| Peripheral vascular disorder due to diabetes mellitus |

| **Arrhythmias** |
|---|
| **Standard Concept Name** |
| Atrial premature complex |
| Paroxysmal ventricular tachycardia |
| Conduction disorder of the heart |
| Ventricular fibrillation |
| Ventricular premature beats |
| Supraventricular tachycardia |
| Cardiac arrest |
| Atrial fibrillation and flutter |
| Typical atrial flutter |
| Left bundle branch block |
| Sinus node dysfunction |
| Bifascicular block |
| Bundle branch block |
| Complete atrioventricular block |
| Premature beats |
| Premature atrial contraction |
| First degree atrioventricular block |
| Cardiac arrhythmia |
| Longstanding persistent atrial fibrillation |
| Ventricular premature complex |
| Paroxysmal atrial fibrillation |
| Left posterior fascicular block |
| **Arrhythmias** |
| **Standard Concept Name** |
| Paroxysmal supraventricular tachycardia |
| Left anterior fascicular block |
| Atrial flutter |
| Right bundle branch block |
| Heart block |
| Permanent atrial fibrillation |
| Atrial fibrillation |
| Aberrant premature complexes |
| Supraventricular premature beats |
| Sick sinus syndrome |
| Cardiac arrest due to cardiac disorder |
| Left bundle branch hemiblock |
| Atypical atrial flutter |
| Long QT syndrome |

Chronic atrial fibrillation

Paroxysmal tachycardia

Fetal dysrhythmia

Persistent atrial fibrillation

[a] an individual can have multiple diagnoses within the same group

[b] detailed hierarchy of the standard concepts can be found in https://athena.ohdsi.org/search-terms/start

### Table S2. Participant counts per antidepressant drug exposure

| Antidepressant Exosures | N cases* |
|---|---|
| sertraline | 56 |
| amitriptyline | ≤ 20 |
| desipramine | 0 |
| duloxetine | 21 |
| escitalopram | 33 |
| nortriptyline | ≤ 20 |
| paroxetine | ≤ 20 |
| trazodone | 28 |
| venlafaxine | 28 |
| vilazodone | ≤ 20 |
| fluoxetine | 33 |
| citalopram | 32 |
| bupropion | 67 |
| doxepin | 0 |
| nefazodone | 0 |
| vortioxetine | ≤ 20 |
| imipramine | 0 |
| amoxapine | 0 |
| **Antidepressant Exosures** | **N cases*** |
| desvenlafaxine | ≤ 20 |
| milnacipran | ≤ 20 |
| mirtazapine | ≤ 20 |
| phenelzine | ≤ 20 |
| protriptyline | 0 |
| 5-hydroxytryptophan | 0 |
| clomipramine | 0 |
| fluvoxamine | 0 |

### Table S3. Antidepressant substances included in each antidepressant group

| Antidepressant Categories | Substances Included |
|---|---|
| SSRIs | sertraline, escitalopram, paroxetine, fluoxetine, citalopram, fluvoxamine |
| TCAs | amitriptyline, desipramine, nortriptyline, doxepin, imipramine, amoxapine, protriptyline, clomipramine |
| SNRIs | venlafaxine, desvenlafaxine, duloxetine, milnacipran |
| SARIs | trazodone, nefazodone, vilazodone |

### Table S4. Effect of number of daily SDANN observations per person on SDANN value

| SDANN association to the Number of Observations | Effect Size | Standard Error | P Value |
|---|---|---|---|
| daily SDANN - Number of observations | -0.003 | 0.013 | 0.84 |
| median SDANN per person - Number of observations | -0.003 | 0.013 | 0.816 |

### Table S5. One-sample Mendelian Randomisation diagnostic test values

| Test | Statistic | P value |
|---|---|---|
| Wald | 5.276 on 14 and 880 DF | <0.001 |
| Wu Hausman | 8.012 | 0.005 |
| Weak Instruments | 7.644 | 0.006 |

#### Table S6. Daily SDANN association with anxiety PRS accounting for antidepressant exposure, BMI, age, sex at birth and the first 10 principal components

| Variable | Estimate | Std. Error | P | 2.5% CI | 97.5% CI |
|---|---|---|---|---|---|
| Anxiety PRS | -0,07737 | 0,02626 | 3,30E-03 | -0,12847 | -0,02627 |
| age | -0,04943 | 0,021729 | 2,30E-02 | -0,09161 | -0,00682 |
| Female sex at birth | -0,38013 | 0,059576 | 2,81E-10 | -0,496 | -0,26413 |
| Baseline BMI | -0,22953 | 0,025889 | 4,18E-18 | -0,2799 | -0,17915 |
| PC1 | 0,019889 | 0,023225 | 3,92E-01 | -0,02531 | 0,065077 |
| PC2 | 0,018389 | 0,024802 | 4,59E-01 | -0,02988 | 0,066637 |
| PC3 | -0,07577 | 0,024459 | 2,01E-03 | -0,12336 | -0,02818 |
| PC4 | -0,00543 | 0,025666 | 8,33E-01 | -0,05539 | 0,044496 |
| PC5 | 0,00022 | 0,025253 | 9,93E-01 | -0,04892 | 0,049358 |
| PC6 | 0,001077 | 0,026589 | 9,68E-01 | -0,05067 | 0,052804 |
| PC7 | 0,014393 | 0,025166 | 5,68E-01 | -0,03459 | 0,063352 |
| PC8 | 0,001598 | 0,025398 | 9,50E-01 | -0,04783 | 0,051013 |
| PC9 | 0,001017 | 0,024055 | 9,66E-01 | -0,04581 | 0,047806 |
| PC10 | -0,06031 | 0,026449 | 2,28E-02 | -0,11176 | -0,00883 |
| Daily antidepressant exposure_yes | -0,0696 | 0,016431 | 2,28E-05 | -0,10184 | -0,03745 |

**Table S7. Anxiety PRS effect on SDANN after excluding people with conditions known to affect heart rate variability**

| | N | Estimate | Std. Error | P | 2.5% CI | 97.5% CI |
|---|---|---|---|---|---|---|
| CVD[a] | 707 | -0,07173 | 0,030516 | 1,90E-02 | -1,31E-01 | -0,01247 |
| MDD[a] | 712 | -0,08591 | 0,030416 | 4,88E-03 | -1,45E-01 | -0,02682 |
| DM[a] | 623 | -0,0755 | 0,02763 | 6,43E-03 | -0,12921 | -0,02178 |

[a]CVD: Cardiovascular Disease; MDD: Major Depressive Disorder; DM: Diabetes Mellitus

**Figure S1. Schematic Representation of One-sample Mendelian Randomization**

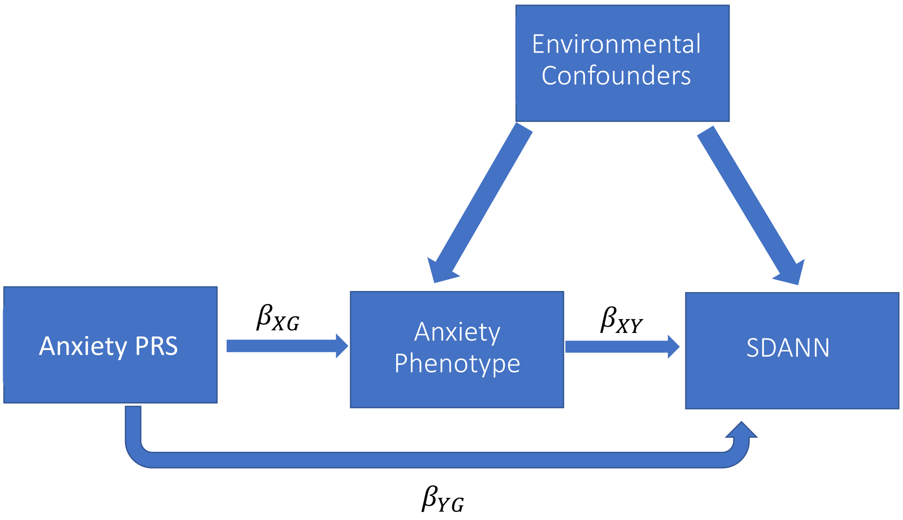


**eFigure1.** The effect $\beta_{XY}$ of our anxiety (exposure X) on the SDANN (outcome Y) equals to $\beta_{XY} = {}^{\beta_{XG}}/_{\beta_{YX}}$, where G is our genetic instrument- the anxiety PRS. It is assumed that the anxiety PRS only affects SDANN through anxiety.

**Figure S2. Distribution of number of daily SDANN observations per individual**

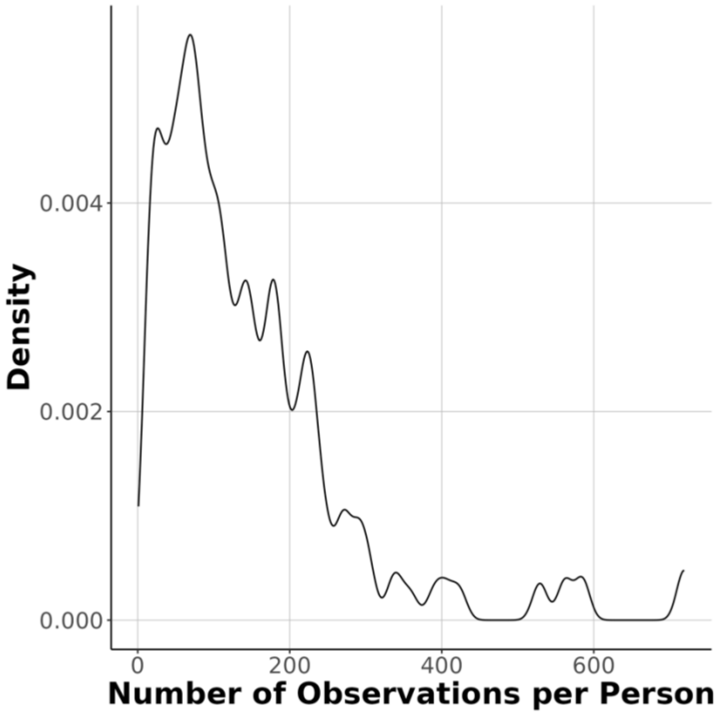

**Figure S3. Effect of age, sex, BMI and anxiety PRS accounting for the first 10 principal components**

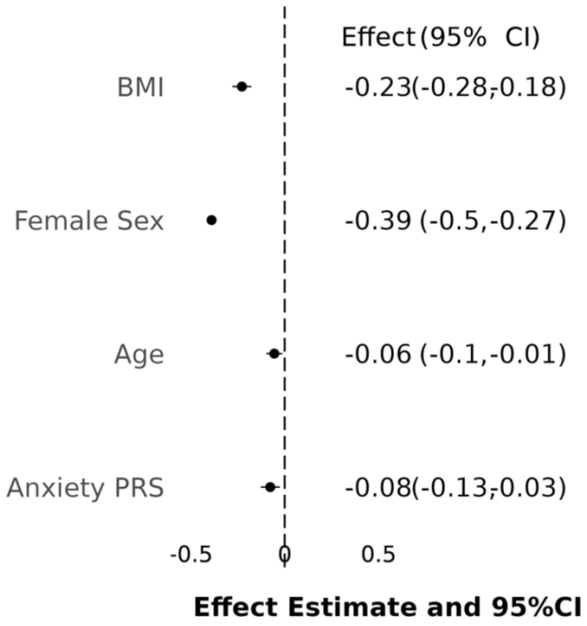
